# Unmet dental needs in children following suspension of school-based oral health services due to COVID-19

**DOI:** 10.1101/2022.05.16.22275165

**Authors:** Ryan Richard Ruff, Tamarinda Barry Godín, Rachel Whittemore, Topaz Murray Small, Nydia Santiago-Galvin, Priyanka Sharma

**Affiliations:** New York University College of Dentistry; New York University School of Global Public Health

## Abstract

**Background:** Dental caries (tooth decay) is the world’s most prevalent noncommunicable disease and can lead to pain, infection, and edentulism. Many children with caries lack access to traditional dental services. School-based caries prevention can increase access to care and reduce health inequities. Disruptions in school-based care due to pandemic control policies may result in children losing access to their primary dental care option.

**Methods:** The CariedAway project was a school-based caries prevention program in operation from 2019-2023 in urban schools with a high proportion of low-income, minority students. Program operations were suspended for two years due to the COVID-19 pandemic. We estimated the prevalence of untreated decay, swelling, fistula, and pulpal involvement in participants at baseline and again after restrictions were lifted.

**Results:** 2998 children between the ages of 5 and 13 years were enrolled and received preventive care prior to pandemic shutdowns, and 1398 (47%) completed their first follow-up observation after two years. At baseline, approximately 30% had untreated caries on any dentition, 11% of children presented with evidence of having received preventive dental sealants, and no participants had swelling, fistula, or pulpal involvement. After 24 months, 12% of participants had swelling fistula, or pulpal involvement that was not treated during the pandemic period.

**Conclusion:** There are considerable unmet dental needs in high-risk children that may be further exacerbated by a lack of access to care during disease outbreaks.

## Introduction

Coined the “silent epidemic” ^1^, the World Health Organization classifies untreated dental caries (cavities) at the most common noncommunicable disease in the world ^2^. Children with caries can experience systemic infection ^3^, lower quality of life ^4^, and reduced academic performance and school attendance ^5^. Inequities in oral disease are a reflection of disparities in access to dental care, as those most at risk have the lowest rates of service utilization ^6^. As a result, multiple federal organizations recommend school-based caries prevention ^7,8^, which can increase access to dental care ^8^ and reduce the burden of disease ^9^.

The COVID-19 pandemic had considerable impact on the availability of dental services, causing a reduction in visits to US dental offices ^10^, a decrease in routine and preventive treatments, and increases in emergency care ^11^. Early interventions to mitigate the spread of COVID-19 before the availability of vaccines included school closings ^12^, which lowered transmission rates ^13^ but also reduced access to school health services ^14,15^. In contrast, school-based health centers that remained open despite school closures observed high encounter rates for preventive health exams ^16^. While not all school caries prevention programs are part of a school-based health center, a substantial proportion of centers provide oral health services in addition to primary care ^17^. To our knowledge there has been no investigation into the impact of the disruption of school dental services due to COVID-19.

CariedAway was a school-based oral health program operating in New York City from 2019 to 2023, providing a range of preventive services including oral examinations, fluoride varnish, silver diamine fluoride, and dental sealants ^18^. Following initial enrollment and treatment, schools participating in CariedAway were closed due to COVID-19 and program operations were suspended from March 2020 to September 2021. Following this suspension, the program completed and primary results were reported ^9^. In this paper, we conduct secondary analysis of the CariedAway data. We describe the baseline oral health of participating children and present data on the prevalence of unmet needs in those who returned following the COVID-19 suspension.

## Methods

CariedAway was a school-based caries prevention program and pragmatic clinical trial that operated from 1 February 2019 to 1 June 2023. The program received IRB approval from the New York University School of Medicine (#i7-00578) and is registered at clinicaltrials.gov (#NCT03442309).

### Settings and Population

Any primary school in New York City with a student population consisting of at least 50% Hispanic/Latino or Black ethnicities and at least 80% receiving free or reduced lunch were eligible to participate. The latter criteria was used as a proxy for low socio-economic status. Schools were further ineligible if they had a preexisting caries prevention program or school based health center providing dental services (SBHC-D) operating within the school.

Inclusion criteria included those with parental informed consent and child assent. Exclusion criteria for individual participants included any child who did not speak English or was enrolled in a special education classroom. While children of any age were eligible for participation in the CariedAway program, analysis only included children ages 5-13.

### Examination and Treatment

All participants in CariedAway received a full visual-tactile dental examination from a standardized dental hygienist or registered nurse while under the supervision of a licensed pediatric dentist. Program participants then received either (a) fluoride varnish (5% NaF, Colgate Prevident) on all teeth with a 38% silver diamine fluoride (Elevate Oral Care Advantage Arrest, 2.24 F-ion mg/dose) solution for asymptomatic cavitated lesions of the posterior dentition and pits and fissures of bicuspids and molars or (b) fluoride varnish on all teeth, glass ionomer sealants (GC Fuji IX, GC America) on pits and fissures of bicuspids and molars, and atraumatic restorations on any asymptomatic cavitated lesions. Both treatments provided primary and secondary prevention. All program participants received treatment at every observation.

### Recruitment and Enrollment

Study investigators first met with the principal and parent coordinator at each school, aTer which a memo of understanding was signed by school administrators to participate in the study. In New York, state approval is required to provided dental care in schools, so applications were submitted to the New York State Department of Health to serve as the sole oral healthcare provider for each school. Following state approval, initial baseline visits were scheduled with each school and consents were distributed to school personnel. During consent delivery, study investigators conducted program promotion in each school, consisting of tabling sessions, informational letters for parents, and presenting at parent-teacher conferences. ATer two weeks, consents were collected and subjects were enrolled and received their assigned treatment.

### Training and Standardization

All clinical research staff, including registered dental hygienists, registered nurses, and dental assistants received a minimum of 70 hours of training during an annual orientation held for the first time in Spring of 2019 and every September thereaTer through the study’s conclusion in Spring 2023. Additional training took place annually during the first week of January (when most elementary schools declined school visits due to conflicts with the academic calendar). Training was didactic and practical, including screening and treatment protocol standardization exercises using intraoral photos and typodonts, as well as through patient interactions with consented students not included in the CariedAway program.

### Outcomes and Diagnostics

We present data for caries prevalence, sealant prevalence, and the prevalence of fistula, pulpal involvement, and swelling. Untreated decay, treated dentition (identified as any tooth presenting with evidence of prior fillings or dental sealants received), and dental sealants were included as dichotomous variables. We also include data on the total number of teeth with untreated decay and the total decayed/missing/filled teeth and tooth surfaces for both deciduous (dmfs/dmT) and permanent (DMFS/DMFT) dentition.

Diagnosis of dental caries was conducted using the International Cares Detection and Assessment System (ICDAS) adapted criteria for epidemiology and clinical research seqngs ^19^. Lesions that were scored as either a 5 (distinct cavity with visible dentin) or 6 (extensive, more than half the surface, distinct cavity with visible dentin) on the ICDAS scale were recorded as caries. Caries was diagnosed at the surface level and analyzed at the individual level (e.g., any participant with any lesion on any surface was classified as having dental caries). Diagnosis of swelling, fistula, or pulpal involvement followed the PUFA index criteria (Presence of visible pulp, Ulceration of the oral mucosa due to root fragments, Fistula, or Abscess). Changes in pulpal status were observed when PUFA criteria were not met at the baseline visit but were met at subsequent observations.

### Statistical Analysis

Sociodemographic information for the enrolled and retained subjects were computed (means, standard deviations, and proportions). We then assessed any sociodemographic variation in baseline untreated caries (present versus not present) or baseline prevalence of dental sealants (present versus not present) using logistic regression. We produced estimates for the prevalence of untreated decay, dental sealants, treated dentition, and the number of decayed, missing, and filled teeth and tooth surfaces for the analytic sample at baseline and follow-up. Finally, we estimated the proportion of subjects with swelling, fistula, or pulpal involvement at each time period and assessed differences using McNemar’s test. Statistical significance was set at p *<*.05. Analysis was conducted using R v1.4.

## Results

A total of 2998 viable subjects were enrolled and treated prior to program suspension due to COVID-19, of which 1398 (47%) completed their follow-up observation after pandemic restrictions were lifted (Table 1). Of the retained participants, 53.9% were female and approximately 64% were either Hispanic or Black race/ethnicities. The average age at baseline amongst retained participants was 6.63 (SD=1.2) and ranged from 5-10 years.

**Table 1:** Sample demographics for enrolled (N=2998) and retained (N=1398) participants.

|  | Enrolled |  | Retained |  |
| --- | --- | --- | --- | --- |
|  | N/Mean | % | N/Mean | % / SD |
| Subjects | 2998 | 100 | 1398 | 100 |
| Female | 1566 | 52.23 | 753 | 53.86 |
| Race |  |  |  |  |
| Hispanic | 1631 | 54.75 | 679 | 48.85 |
| Black | 558 | 18.73 | 208 | 14.96 |
| White | 75 | 2.52 | 29 | 2.09 |
| Asian | 44 | 1.48 | 24 | 1.73 |
| Multiple | 58 | 1.95 | 20 | 1.44 |
| Other | 41 | 1.38 | 11 | 0.79 |
| DK/Missing | 572 | 19.20 | 419 | 30.14 |
| Age at baseline | 6.63<br>(mean) | 1.24<br>(SD) | 6.63<br>(mean) | 1.21<br>(SD) |

For logistic regression models predicting untreated decay or the presence of sealants at baseline (Table 2), Black participants were less likely to have sealants compared to Hispanic/Latino participants (OR = 0.54, 95% CI = 0.30, 0.96). The odds of having sealants increased with age at baseline (OR = 1.85, 95% CI = 1.59, 2.15), as did the odds of having untreated decay (OR = 1.11, 95% CI = 1.01, 1.22). There were no significant differences in the odds of decay or sealants by sex.

**Table 2:** Odds of untreated decay and sealants at baseline for sociodemographic groups (N=1398)

|  | Untreated decay |  | Sealant present |  |
| --- | --- | --- | --- | --- |
|  | OR | 95% CI | OR | 95% CI |
| Race |  |  |  |  |
| Black | 1.29 | 0.92, 1.80 | 0.54 | 0.30, 0.96 |
| White | 1.47 | 0.67, 3.22 | 1.41 | 0.47, 4.27 |
| Asian | 1.42 | 0.60, 3.39 | 0.79 | 0.18, 3.56 |
| Multiple | 0.87 | 0.31, 2.44 | 0.35 | 0.44, 2.71 |
| Other | 0.59 | 0.13, 2.75 | 0.6 | 0.73, 4.85 |
| Unreported | 1.31 | 1.00, 1.70 | 0.98 | 0.67, 1.44 |
| Age | 1.11 | 1.01, 1.22 | 1.85 | 1.59, 2.15 |
| Females | 0.89 | 0.71, 1.13 | 1.15 | 0.81, 1.62 |

*Notes: Logistic regression results for outcomes of untreated decay and presence of sealants at baseline. Models included all explanatory variables for race/ethnicity, age, and sex. Reference groups include Hispanic/Latino for race/ethnicity and males for sex*

Of retained participants, 413 participants presented at baseline with untreated decay (29.5%, 95% CI = 27.21, 32.0). After receiving treatment for existing caries and prevention for sound teeth, 380 participants (27.2%, 95% CI = 24.91, 29.58) had untreated decay again at their post-pandemic observation (Table 3). Approximately 21% of participants had a filling when they received their baseline observation, and only 11% had preexisting sealants. At follow-up, reflecting the treatment received through CariedAway, 41% of participants had sealants present. The remaining participants received silver diamine fluoride, as part of the original clinical protocol. The average decayed, missing, or filled index at follow-up was 1.1 (SD=1.8) for deciduous teeth, 3.1 (SD=6.0) for deciduous surfaces, 0.1 (SD=0.4) for permanent teeth, and 0.2 (0.8) for permanent surfaces.

**Table 3:** Oral health status at baseline and follow-up (N=1398)

|  | Baseline |  | Follow-up |  |
| --- | --- | --- | --- | --- |
|  | N | % (total) | N | % (total) |
| Untreated decay, overall | 413 | 29.54 | 380 | 27.18 |
| Decay, occlusal first molar | 26 | 1.86 | 41 | 2.93 |
| Decay, deciduous | 401 | 28.68 | 343 | 24.54 |
| Decay, permanent | 30 | 2.15 | 58 | 4.15 |
| Total decayed teeth |  |  |  |  |
| 0 | 985 | 70.46 | 1018 | 72.82 |
| 1-2 | 269 | 19.24 | 294 | 21.03 |
| 3-5 | 121 | 8.66 | 71 | 5.08 |
| 6-8 | 22 | 1.57 | 13 | 0.93 |
| 8+ | 1 | 0.07 | 4 | 0.28 |
| Decay by age |  |  |  |  |
| 5 | 78 | 18.89 | * | * |
| 6 | 106 | 25.67 |  |  |
| 7 | 102 | 24.70 |  |  |
| 8 | 103 | 24.94 |  |  |
| 9 | 23 | 5.57 |  |  |
| 10 | 1 | 0.24 |  |  |
| Decay by race/ethnicity |  |  |  |  |
| Hispanic | 183 | 44.63 | 219 | 58.09 |
| Black | 67 | 16.34 | 87 | 23.08 |
| White | 10 | 2.44 | 5 | 1.33 |
| Asian | 8 | 1.95 | 6 | 1.59 |
| Other | 2 | 0.49 | 10 | 2.65 |
| Multiple | 5 | 1.22 | 1 | 0.27 |
| Unreported | 135 | 32.93 | 49 | 13.00 |
| Baseline treated dentition | 374 | 26.75 | NA | NA |
| Sealant prevalence | 156 | 11.16 | 568 | 40.63 |
| Filling prevalence | 288 | 20.6 | 338 | 24.18 |
| Sealant by race |  |  |  |  |
| Hispanic | 83 | 53.55 | 357 | 63.30 |
| Black | 15 | 9.68 | 102 | 18.09 |
| White | 4 | 2.58 | 11 | 1.95 |
| Asian | 2 | 1.29 | 11 | 1.95 |
| Other | 1 | 0.65 | 5 | 0.89 |
| Multiple | 1 | 0.65 | 11 | 1.95 |
| Unreported | 49 | 31.61 | 67 | 11.88 |
|  | X | SD | X | SD |
| DMFT | 0.54 | 0.3 | 0.12 | 0.43 |
| DMFS | 0.79 | 0.49 | 0.19 | 0.78 |
| dmft | 1.24 | 2.04 | 1.06 | 1.82 |
| dmfs | 3.42 | 6.43 | 3.10 | 5.97 |

Notes: DMFT/dmft is the decayed, missing, and filled tooth index for permanent and deciduous teeth, respectively. DMFS/dmfs is the decayed, missing, and filled surfaces for permanent and deciduous teeth, respectively.
\* Age was only assessed at baseline

There were no participants at baseline with fistula, pulpal involvement, or swelling (Table 4). At follow-up, 171 participants (12.2%) presented with either fistula, pulpal involvement, or swelling (p < .00001). This consisted of 61 participants (4.4) having fistula (p < .00001), 138 (9.9%) having pulpal involvement (p < .00001), and 6 (0.4%) having swelling (p = 0.014).

**Table 4:**
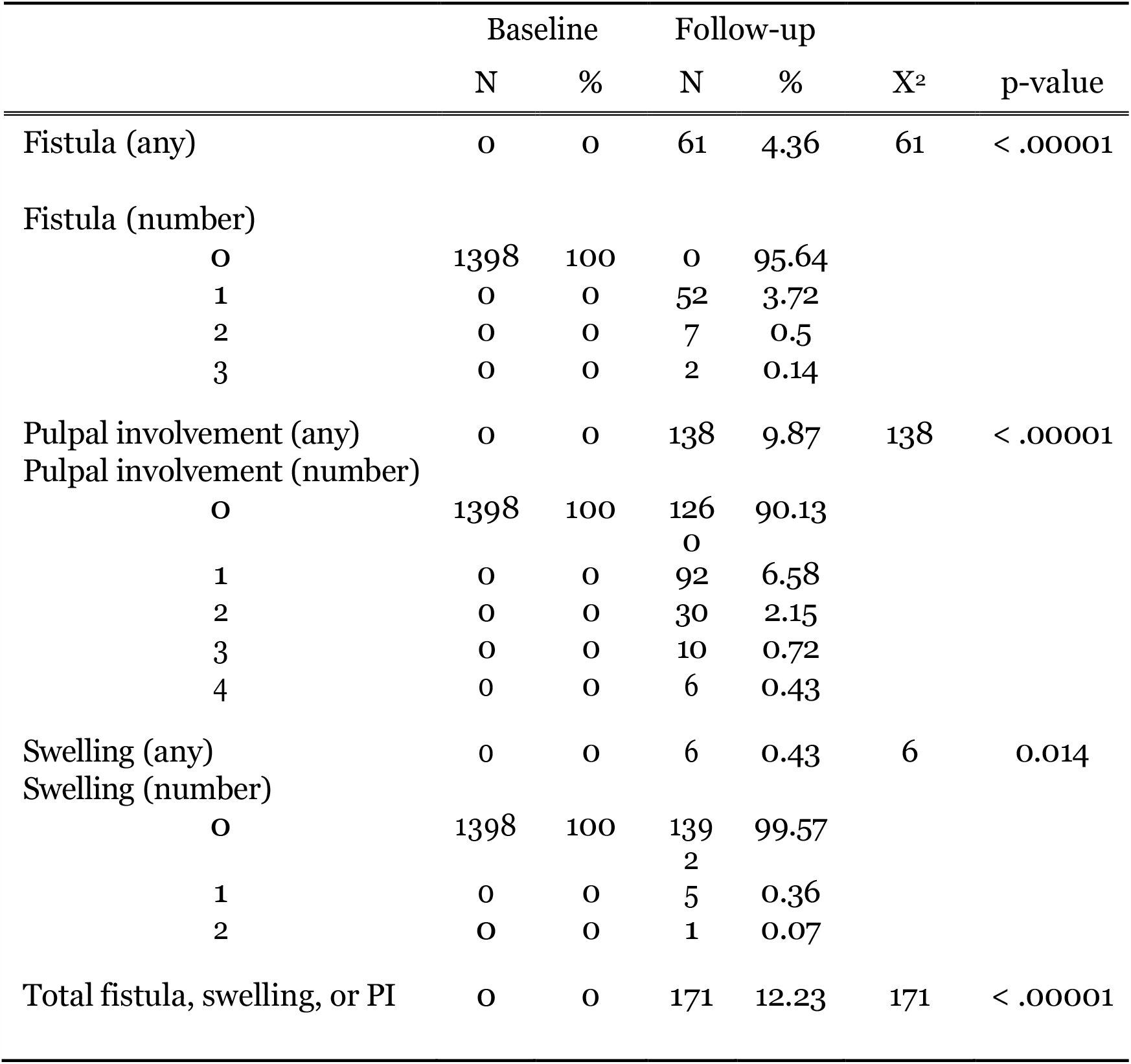
Prevalence of fistula, pulpal involvement, and swelling at baseline and follow-up (N=1398)

## Discussion

School-based caries prevention programs are effective and cost-efficient methods to increase access to care for vulnerable, traditionally underserved populations ^7,8,20^ and can significantly reduce the risk of disease ^21^. However, no preventive therapy is 100% effective. Prior studies into school caries prevention conclude that approximately 20-30% of participating students go on to develop new dental caries ^9,21^. As a result, continued care for these children is required to ensure that oral diseases are controlled before they progress to more serious infection. Results from secondary data analysis of the CariedAway school-based caries prevention program suggest that children who experienced new dental caries or developed swelling, fistula, or pulpal involvement did not seek outside care from dental offices when program services were suspended due to the COVID-19 pandemic.

The observed baseline prevalence of untreated caries and dental sealants indicates a lack of access to, or utilization of, office-based dental services and is consistent with the extant literature. In a similar assessment of low-income children participating in a school-based sealant program in Massachuse’s, approximately one third had untreated caries despite living in close proximity to community dental clinics and nearly two-thirds of children had no clinical indication of prior treatment with sealants on permanent dentition ^22^. The fact that dental fillings were observed in three times as many CariedAway subjects as sealants suggest that the majority of outside care, when provided, was for caries management and not prevention. As a result, school-based health programs likely serve as the predominant source of caries prevention for this population.

In New York City, dental offices were closed in June 2020 except for emergency care and all school-based health programs were suspended from March 2020 through August 2021. While traditional offices later reopened with stringent infection control policies in place, dental care access during the pandemic was significantly reduced ^23^. Although there were no significant differences in untreated decay between baseline and follow-up, the treatment protocols in CariedAway included both primary (prevention of new disease) and secondary (arrest of existing disease) prevention. As a result, all untreated caries at baseline were arrested using either silver diamine fluoride or atraumatic restorations. This means that approximately 30% of participants experienced new caries during the pandemic period that were not treated. Similarly, approximately 12% of subjects returning to the program after the pandemic had fistula, swelling, or pulpal involvement that were not treated while the project was suspended. Research suggests that children were more likely to have poor oral health and a lower utilization of dental visits during the COVID-19 period, specifically preventive ones ^24^, and the largest decline in school health services due to the pandemic were reported for dental screenings and dental services ^25^. In contrast, those school health services that remained open for clinical care throughout COVID-19 saw increased annual preventive health exams and were a critical source of primary care to vulnerable populations ^16^.

Actual versus perceived availability of affordable dental health options may further exacerbate oral disease inequities in urban school children. Approximately 4600 dental providers were previously listed that operated in New York, treated children, and accepted both Medicaid and the Children’s Health Insurance Program. However, 90% of these providers were located in a small number of counties, and a substantial proportion of listings for providers were invalid ^26^. As a result, children from low-income households may not have the support necessary to treat urgent needs outside of school-based dental care.

### Implications for School Health

Suspension of school-based health services due to increased restrictions to control the spread of infectious disease, such as those implemented in response to COVID-19, can potentially result in a lack of adequate dental care for high-need children. Concerns with school-based dental operations during COVID-19 stemmed from the risks of aerosolization, which can increase the airborne transmission of viruses. While this concern is justified, the silver diamine fluoride implemented in CariedAway is non-aerosol generating ^27,28^ and is recommended as part of a Safer Aerosol-Free Emergent Dentistry (SAFER) approach to dental care ^29^. Although recommendations for preparing for future pandemics focus largely on virology, transmission models, and rapid vaccine development ^30^, attention should also be given to minimize disruption in community health services during pandemic periods. Establishing emergency protocols for school-health services that can safely treat children who otherwise lack access is critical to ensure continuity of care.

## Data Availability

CariedAway is an active trial. Data requests will be reviewed and may be available upon approval.

## Author contributions

RRR was a principal investigator and conceived of and designed the study. TBG served as the supervising dentist, directed clinical activities, and oversaw all data collection. RW, TM, NSG, and PS were involved in the conduct and management of CariedAway, including study coordination, enrollment, community engagement, provision of treatments, and data collection. RRR wrote the manuscript. All authors critically reviewed the manuscript and provided edits. All authors read and approved the final manuscript.

## Availability of Data and Materials

The datasets generated and/or analyzed during the current study are not publicly available, but are available from the corresponding author upon reasonable request.

